# Prevalence of Physical Activity Initiatives in Australian Primary Schools: A Cross-Sectional Survey

**DOI:** 10.1101/2024.11.20.24317670

**Authors:** Kate M O’Brien, Jessica Bell, Luke Wolfenden, Nicole Nathan, Serene Yoong, Adrian Bauman, Christophe Lecathelinais, Lucy Leigh, Rebecca K Hodder

## Abstract

**Introduction:** Schools play a significant role in influencing children’s physical activity and sedentary behaviours and are a key recommended setting for physical activity initiative globally. To achieve population-wide health benefits, they must be guideline-informed and widely adopted. However, evidence on their implementation in Australian primary schools is limited.

**Objective:** To assess the reported implementation of recommended physical activity initiatives in Australian primary schools and explore their associations with school characteristics.

**Methods:** A cross-sectional study surveyed a nationally representative sample of Australian primary school principals (August 2022-October 2023). Principals reported on 33 physical activity initiatives categorised by delivery context: in the classroom; outside the classroom and during break times; outside of school outside or involving families; or other. Prevalence estimates were weighted to the Australian school population, and logistic regression models assessed associations with school characteristics.

**Results:** A total of 669 schools participated, with 360 principals completing the survey. Reported implementation rates varied from 4% to 98%. The most frequently implemented initiative in classrooms was *Physical activity units of work in PDHPE/HPE curriculum across all year groups* (98%); outside the classroom/break times was *School infrastructure that supports physical activity during breaks everyday (e*.*g. play areas)* (96%); and outside of school/involving families was *School provides end of trip facilities to encourage active school travel* (75%). Implementation of nine initiatives was associated with school size (n=6), geographic location (n=4), or socio-economic status (n=1).

**Conclusion:** This first national study provides critical information on current implementation levels individual physical activity initiatives in Australian primary schools and highlights where policy and practice investment in implementation support is required.

## Introduction

Australian and international guidelines recommend that children five years and older do at least an average of 60 minutes per day of moderate-to-vigorous physical activity (MVPA) across the week and limit the amount of time spent being sedentary, particularly recreational screen time^1–3^ State-based surveys included in the Active Healthy Kids Australia 2022 report card indicated that, according to self-reports from students and parents, between 19% and 67% of primary school children meet the physical activity guidelines for daily MVPA.^4^ This significant variation could be attributed to measurement bias.^5^ However, the proportion of children meeting these guidelines based on device-measured physical activity is also low^6^. Given the significant burden of physical inactivity, Australia’s National Preventive Health Strategy 2021-2023^7^ and the National Obesity Strategy 2022-2032^8^ include goals to reduce the prevalence of physical inactivity amongst children, adolescents and adults by at least 15% by 2030.^7^

Schools are crucial in shaping children’s physical activity and sedentary behaviours.^3^ Schools reach many children over extended periods, creating a valuable opportunity to deliver quality physical activity education, promote active school days and reduce chronic diseases on a population scale.^3^ Consequently, substantial research investment has focused on identifying effective school-based physical activity initiatives. Systematic reviews show that these initiatives generally increase children’s MVPA.^3, 9, 10^ For example, a recent Cochrane systematic review of randomised controlled trials (RCTs) found that school-based physical activity initiatives implemented for at least 12 weeks improved MVPA in children by a mean difference of 0.73 minutes per day (95% confidence interval (CI) 0.16 to 1.30; 33 trials; 20,614 participants).^3^

To ensure children benefit from a physical activity promoting school environment, governments and health organisations globally provide recommendations for specific physical activity initiatives in schools.^11–14^ For example, the World Health Organisation (WHO) recommends a comprehensive physical activity policy in schools, which includes quality PE, promoting active school travel, offering physical activity opportunities during breaks, and integrating activity into classroom lessons.^11^ In Australia, primary schools are encouraged to adopt a whole-of-school approach to physical activity alongside mandatory PE.^15^ While there is no nationwide mandatory policy, various states have established specific policies outlining recommended initiatives,^13, 16–19^ such as Victoria’s Active Schools toolkit^19^ and New South Wales (NSW) Live Life Well @ School (LLW@S) program.^13^ These programs recommend delivering quality PE, fostering fundamental movement skills, creating supportive environments for physical activities (e.g. sports equipment, playground activities), promoting active classrooms (e.g. active breaks), and encouraging active school travel. To maximise child health benefits and achieve public health objectives, these policies must be implemented widely and aligned with established guidelines.^20^

Despite significant investment and guideline recommendations, there is no national evidence of implementation of a comprehensive range of physical activity initiatives in Australian primary schools. Existing data primarily focuses on specific states or limited interventions. For instance, a 2013 study of 303 NSW primary schools reported the implementation of five initiatives, ranging from 59% of schools integrating physical activity into other subjects to 82% informing parents about physical activity.^21^ A subsequent NSW study of 115 schools (2016-2021) found that 59% met the quality PE requirement of 150 minutes per week, while 48% used classroom energisers and 75% included active homework.^22^ These findings, while informative, highlight the need for more comprehensive national data on the range of physical activity initiatives implemented in Australian primary schools.

Examining the associations between physical activity initiatives implemented in primary schools and characteristics like school size, remoteness, and socio-economic status is essential for tailoring effective interventions. Understanding these characteristics allows for targeted support for schools facing unique challenges, ensuring that resources are allocated where they are most needed. For example, schools in disadvantaged areas may encounter barriers to implementing programs, so addressing these disparities is important for achieving equitable health outcomes for all children. National data on implementation across these characteristics can inform evidence-based policies and enhance program engagement, fostering a culture of health and wellness in schools. Despite previous research indicating variations in the implementation of physical activity initiatives based on school size, geographic location, and socio-economic status,^23^ there is currently no national data available on this issue.

Given the limited evidence, a national cross-sectional study was conducted to determine the current implementation of a broad range of recommended physical activity initiatives in Australian primary schools and to determine whether implementation is associated with school size, remoteness, or socio-economic status.

## Methods

### Ethical approval

Approval to conduct the study was received from the University of Newcastle Human Research Ethics Committee (approval no. H-2021-0045) and 25 out of 32 Australian school jurisdictions nationally (Appendix A).

### Study design and setting

A cross-sectional study was conducted between August 2022 and October 2023, using a nationally representative sample of Australian primary school principals from all Australian states and territories.

### Sample

#### Schools

Schools from all Australian states and territories and all education sectors (government, catholic and independent schools) with primary school enrolments were eligible for participation. Non-mainstream schools (e.g. those for special purposes such as students with intellectual disabilities); language or mature-age schools, preschools, schools providing short-term education, or schools within school jurisdictions that did not approve the research were not eligible.

An equal probability stratified sampling methodology was used by the Australian Council for Education Research (ACER) to achieve a nationally representative random sample of 4500 Australian primary schools. Schools were stratified according to state/territory, education sector (government, catholic, and independent schools), remoteness (very remote, remote, outer regional, inner regional, urban areas), socio-economic status (using Socio-Economic Indexes for Areas (SEIFA) Index of Education (IEO) National Decile) and school size (total enrolment quartiles).

### Participants

Principals of all selected schools, or their nominated delegates, were eligible to participate in the study.

### Recruitment and data collection procedures

School principals of eligible schools were sent an information pack to a publicly available school email address to invite them to participate inclusive of an information statement, and an online consent form through which they could indicate their preference to complete the survey online (via the Research Electronic Data Capture platform (Redcap)^24^ or via a Computer Assisted Telephone Interview (CATI). Principals who opted to complete the survey online were sent a unique REDCap survey link. Those choosing to complete the survey via CATI were telephoned by a trained interviewer who entered responses into the REDCap platform; both methods took approximately 30 minutes to complete.

One week following the study invitation, non-responding principals were prompted by email, and one week later non-responding principals received up to 10 telephone prompts to complete the survey by trained interviewers. The protocol for prompting non-responding principals was amended for three school jurisdictions to reduce or omit telephone prompts. All eligible schools were invited to participate, and the prompting of schools continued until the target quota for each state or territory was achieved.

### Measures

#### School characteristics

Data regarding school characteristics were sourced from the ACER database of all Australian schools by states and territories, and supplemented by data from the publicly available Australian Curriculum, Assessment and Reporting Authority (ACARA)^25^ and included: state/territory; education sector; jurisdiction or Archdiocese/Diocese; Australian Statistical Geography Standard (ASGS) remoteness category; socio-economic status; school size (total enrolment); and total number of student enrolment and proportion who are Aboriginal and/or Torres Strait Islander.

#### Participant characteristics

Principals reported their current role and duration worked in that role via the principal survey.

#### Implementation of physical activity initiatives

To reduce participant burden, schools were randomised (using stratified randomisation in SAS Version 9.4^26^ by state/territory and sector) to answer survey questions regarding the physical activity initiatives within their school. Survey items asked principals to report whether their school was currently implementing 33 individual physical activity initiatives. Physical activity initiatives previously adopted in school-based programs globally were identified via a series of recent high-quality systematic reviews of school-based physical activity programs^3, 27^ that align with recommendations of Australian and global policies and guidelines.^16, 28–34^ These initiatives were categorised into four opportunities for physical activity according to Australian best practice recommendations for school-based physical activity strategies: physical activity in the classroom (13), physical activity outside the classroom and during break times (10), physical activity outside of school outside or involving families (6), and other (4) (see Appendix B for physical activity initiatives definitions). Existing validated or published survey items were mapped to each physical activity initiative, or where not available, developed by the research team (see Appendix C for Principal survey). The principal survey was piloted with research staff with teaching experience, stakeholders within the sector and school principals following ethics approval and modifications made prior to implementation to improve interpretation and data quality.

### Sample size

A sample of 350 schools was estimated to be sufficient to generate a weighted national prevalence of physical activity initiatives implementation with a precision of 5.2%.

### Statistical analysis

#### School and participant characteristics

School postcodes were used to classify schools using the national SEIFA 2016 index of Relative Socio-economic Disadvantage (IRSD) which focuses on relative socio-economic disadvantage.^35^ Descriptive statistics were used to summarise school and principal characteristics and participation rates. Schools that participated in the survey were compared to non-participants using chi-square analyses on characteristics (state/territory, school type, rurality, SES, school size).

#### Implementation of physical activity initiatives

The definitions for implementing physical activity initiatives were developed through consensus among researchers, stakeholders, and practitioners with expertise in this area of research. For example, the initiative *‘Physical activity units of work in Personal Development, Health and Physical Education (PDHPE)/HPE curriculum across all year groups’* was assessed using the survey question “Do classes at your school integrate physical activity into Key Learning Areas other than PDHPE/HPE?”, the four response options provided were categorised as either implemented (response options: “yes, all classes” or “yes, some classes”) or not implemented (response options: “no” or “unsure”).

The prevalence estimates of implementation of each physical activity initiative were adjusted by raking the survey weights iteratively to align sample distributions with the Australian school population considering the stratification variables jurisdiction, sector, SEIFA, ASGS remoteness category, and school size quartile), and mitigating non-response bias using SAS Version 9.4.^26^ Integrated and tailored weight trimming during raking iterations were applied, reducing excessive weights while maintaining alignment with population controls to generate the weight prevalence estimates for each physical activity initiative.

#### Association between implementation and school characteristics

Weighted simple logistic regression models were conducted to examine the associations between the implementation of physical activity initiatives and individual school characteristics: school size (small <300/large >=300), remoteness (urban: major city/rural: inner regional, outer regional, remote) and socio-economic status (most disadvantaged/least disadvantaged). Odds ratios (OR) and 95% CI were calculated for each model. Statistical analyses were performed using SAS Version 9.4.^26^ Statistical significance was set at an alpha level of 0.05.

## Results

### School and principal characteristics

There were 7793 Australian schools with primary school enrolment identified, of which 514 were deemed ineligible (e.g. non-mainstream schools) prior to sampling. Of the 4500 schools identified via the stratified random sampling method, 1064 (24%) schools were classified as out of scope (see Appendix D for reasons for out of scope) leaving 3436 eligible schools in the sampling frame. Of these 3303 primary schools were invited to participate in the survey which was conducted between 9 August 2022 and 20 October 2023 (133 schools not invited due to meeting quota for that state). In total, 669 schools completed the survey (via telephone n=57, online n=612; 20% participation rate) and 460 (14%) declined.

Of the 669, 360 schools completed the physical activity survey items. The characteristics of the schools and principals are reported in Table 1. The majority of participating schools were from the government sector (66%), located outside urban areas (61%), situated in the most disadvantaged areas (63%), and were small schools (<300 students; 66%). School enrolments ranged from 2 to 1569 students (mean 289) with a mean of 12% indigenous enrolments. Most participants were school Principals (88%) and had on average worked for 5 years. The demographic profile of the participating schools broadly mirrored the overall distribution of primary schools in Australia, except for NSW, where government school research approval was not extended. Additionally, a higher proportion of small schools from disadvantaged areas participated in the survey compared to their representation in the national primary school demographic. There were significant differences in the school characteristics of participating versus non-participating eligible schools for state (p<0.001), sector (0.008), location (p<0.001), SES (p=0.005), and school size (p<0.001) (Table 1).

**Table 1.** Schools and principal characteristics.

| Characteristics | N=360 schools<br>n (%) | N=2402 schools<br>n (%) | Eligible Australian<br>schools<sup>a</sup><br>(%) |
| --- | --- | --- | --- |
| **State/territory** |  |  |  |
| ACT | 5 (1.39) | 30 (1.25) | 1.39 |
| NSW | 36 (10.00) | 663 (27.60) | 31.79 |
| NT | 15 (4.17) | 66 (2.75) | 2.16 |
| QLD | 118 (32.78) | 563 (23.44) | 18.60 |
| SA | 30 (8.33) | 183 (7.62) | 7.88 |
| TAS | 15 (4.17) | 69 (2.87) | 2.77 |
| VIC | 93 (25.83) | 558 (23.23) | 23.72 |
| WA | 48 (13.33) | 270 (11.24) | 11.69 |
| **Sector** |  |  |  |
| Government | 238 (66.11) | 1744 (72.61) | 69.29 |
| Catholic | 70 (19.44) | 355 (14.78) | 17.85 |
| Independent | 52 (14.44) | 303 (12.61) | 12.85 |
| **Remoteness** |  |  |  |
| Urban<sup>b</sup> | 141 (39.17) | 1168 (48.63) | 51.35 |
| Rural<sup>c</sup> | 219 (60.83) | 1234 (51.37) | 48.65 |
| **SES** |  |  |  |
| Most Disadvantaged | 226 (62.78) | 1346 (56.04) | 52.83 |
| Least Disadvantaged | 134 (37.22) | 1056 (43.96) | 47.17 |
| **School size** |  |  |  |
| Small (<300 students) | 237 (65.83) | 1295 (53.91) | 52.67 |
| Large (≥300 students) | 123 (34.17) | 1107 (46.09) | 47.33 |
| **Total enrolment**, mean (SD), range | 289.04 (302.98), 2 to 1569 | 353.71 (353.61), 2 to 3345 | 366.21 (380.78), 2 to 4726 |
| **Indigenous Enrolments**, mean % (SD) | 11.55 (19.76) | 11.68 (19.62) | 10.67 (18.55) |
| **Staff Role who completed the survey** |  |  |  |
| Principal<sup>d</sup> | 315 (87.50) | - | - |
| Deputy/Assistant Principal | 22 (6.11) | - | - |
| Head of school/Campus/Operations | 5 (1.39) | - | - |
| Teacher/Coordinator<sup>e</sup> | 12 (3.33) | - | - |
| Other role<sup>f</sup> | 6 (1.67) | - | - |
| **How long been in this role** |  |  |  |
| 12 months or more, range in years | 282 (78.33), 1 to 35 | - | - |
| Less than 12 months | 78 (21.67) | - | - |

Reported as n (%), unless otherwise stated; ACT: Australian Capital Territory; QLD: Queensland; NSW: New South Wales; NT: Northern Territory; SA: South Australia; SD: standard deviation; SES: Socio-Economic Status; TAS: Tasmania; VIC: Victoria; WA: Western Australia; <sup>a</sup>Data from the publicly available Australian Curriculum, Assessment and Reporting Authority; <sup>b</sup>Includes Urban areas; <sup>c</sup>Includes inner regional, outer regional, remote and very remote; <sup>d</sup>Executive principal, principal, acting principal; <sup>e</sup>Lead teacher, classroom teacher, health and wellbeing teacher, coordinator; <sup>f</sup>Administration officer, business manager, school counsellor/social worker, school nurse.

### Weighted prevalence of implementation of physical activity initiatives

Table 2 provides a detailed description of the weighted prevalence of the 33 implemented physical activity initiatives across the four opportunities for physical activity.

**Table 2.**
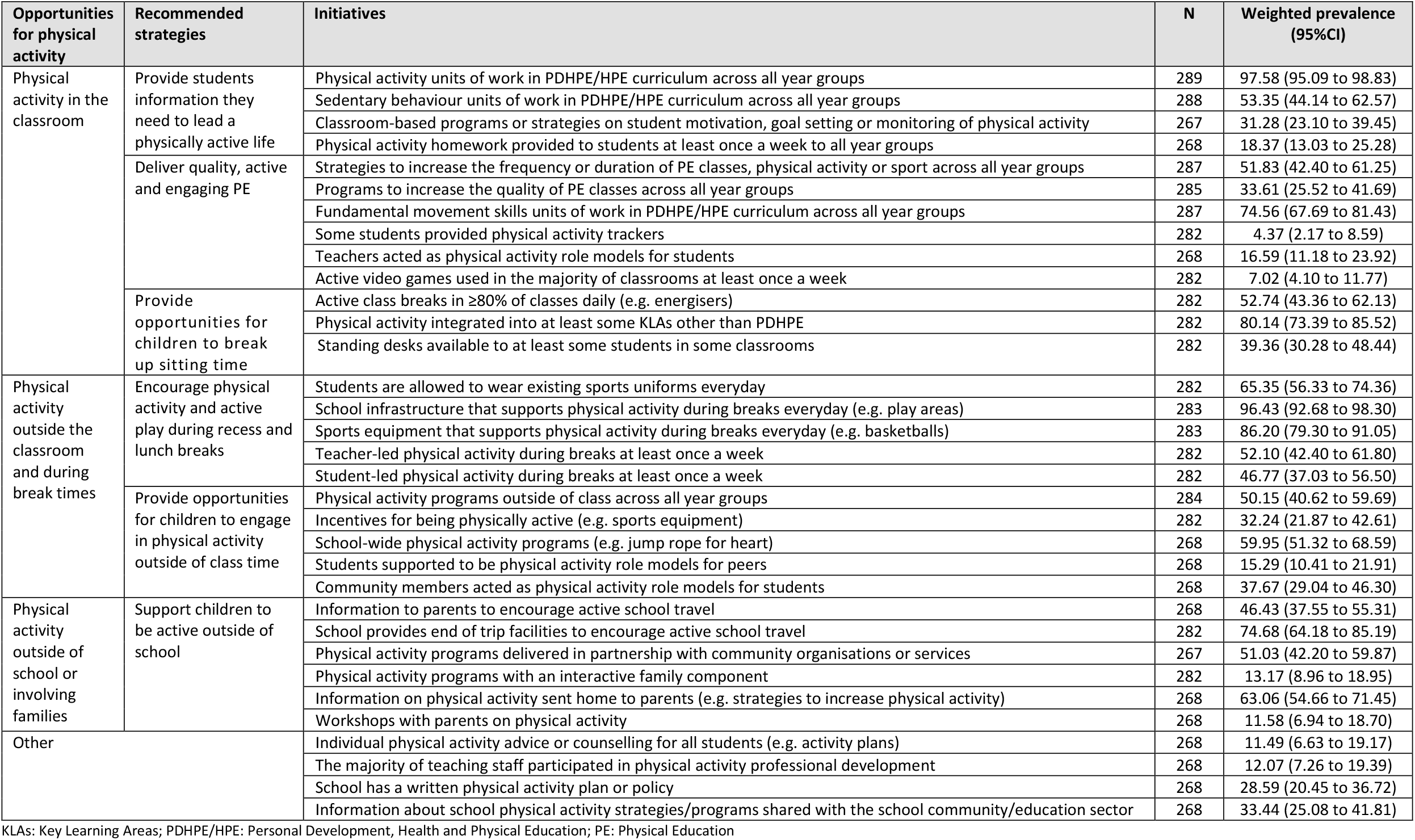
Weighted prevalence of implementation of the 33 physical activity initiatives.

### Physical activity in the classroom

For physical activity in the classroom, the weighted prevalence varied from 4% for *‘Some students provided physical activity trackers’* to 98% for *‘Physical activity units of work in PDHPE/HPE curriculum across all year groups’*. Other initiatives implemented in the majority of schools were *‘Physical activity integrated into at least some Key Learning Areas other than PDHPE’* (80%), *‘Fundamental movement skills units of work in PDHPE/HPE curriculum across all year groups’* (75%), ‘*Sedentary behaviour units of work in PDHPE/HPE curriculum across all year groups’* and *‘Active class breaks in ≥80% of classes daily (e*.*g. energisers)’* (both 53%) and *‘Strategies to increase the frequency or duration of PE classes, physical activity, or sport across all year groups’* (52%). For this item participants also were asked to provide a brief description of the strategies used, some of the most reported strategies were additional minutes per day for fitness, extra break time for the whole school and using Sporting Schools funding to employ specialist coaches for additional sports and activities.

### Physical activity outside the classroom and during break times

When examining physical activity outside of the classroom and during break times, the weighted prevalence showed a range from 15% for *‘Students supported to be physical activity role models for peers’* to 96% for *‘School infrastructure that supports physical activity during breaks everyday (e*.*g. play areas)’. ‘Students are allowed to wear existing sports uniforms everyday’* and *‘Sports equipment that supports physical activity during breaks everyday (e*.*g. basketballs)’* were also widely used (65% and 86%, respectively). Furthermore, *‘School-wide physical activity programs (e*.*g. jump rope for heart)’, ‘Teacher-led physical activity during breaks at least once a week’* and *‘Physical activity programs outside of class across all year groups’* were adopted by 60%, 52% and 50% of schools, respectively.

Physical activity outside of school or involving families

The weighted prevalence of physical activity outside of school or involving families ranged from 12% for *‘Workshops with parents on physical activity’* to 75% for *‘School provides end of trip facilities to encourage active school travel’*. The other physical activity initiatives implemented in the majority of schools involving families were disseminating *‘Information on physical activity sent home to parents (e*.*g. strategies to increase physical activity’* (63%) and *‘Physical activity programs delivered in partnership with community organisations or services’* (51%).

### Other

Of the four physical activity initiatives categorised as other, none were implemented in over 50% of schools: *‘Individual physical activity advice or counselling for all students (e*.*g. activity plans)’* (11%), ‘*The majority of teaching staff participated in physical activity professional development’* (12%), *‘School has a written physical activity plan or policy’* (29%) and *‘Information about school physical activity strategies/programs shared with the school community/education sector’* (33%).

### Association between school characteristics and implementation of physical activity initiatives

Associations between school size, location or socio-economic status were found for nine of the 33 initiatives summarised below (Appendix E). Of the 33, significant findings were found for nine of the physical activity initiatives for school size (6 initiatives), geographic location (4 initiatives) and socio-economic status (1 initiative).

### School size

Compared to small schools (<300 enrolments), large schools had higher odds of implementing four initiatives. Large schools had higher odds of implementing *‘Sedentary behaviour units of work in PDHPE/HPE curriculum across all year groups’* (63% vs. 45%; OR 2.08 (95%CI 1.02 to 4.35)), *‘Classroom-based programs or strategies on student motivation, goal setting or monitoring of physical activity’* (41% vs. 23%; OR 2.27 (95%CI 1.08 to 5.00)), *‘Incentives for being physically active (e*.*g. sports equipment)’* (43% vs. 23%; OR 2.56 (95%CI 1.08 to 6.25)), and *‘Information on physical activity sent home to parents (e*.*g. strategies to increase physical activity)’* (75% vs. 52%; OR 2.70 (95%CI 1.19 to 6.25)) compared to small schools.

Compared to large schools, small schools had higher odds of implementing two initiatives. Small schools had higher odds of implementing *‘Programs to increase the quality of PE classes across all year groups’* (42% vs. 24%; OR 2.23 (95%CI 1.05 to 4.73)) and *‘Physical activity programs with an interactive family component’* (19% vs. 7%; OR 3.30 (95%CI 1.23 to 8.86)) compared to large schools.

### Geographic location

Compared to schools in rural areas, schools in urban areas had higher odds of implementing four initiatives. Schools in urban areas had higher odds of implementing ‘*Physical activity programs outside of class across all year groups’* (59% vs. 41%; OR 2.13 (95%CI 1.04 to 4.35)), *‘Incentives for being physically active (e*.*g. sports equipment)’* (42% vs. 22%; OR 2.70 (95%CI 1.15 to 6.25)), *‘Information to parents to encourage active school travel’* (60% vs. 32%; OR 3.23 (95%CI 1.49 to 6.67)), and *‘Information on physical activity sent home to parents (e*.*g. strategies to increase physical activity)’* (73% vs. 52%; OR 2.5 (95%CI 1.12 to 5.56)) compared to schools in rural areas.

### Socio-economic status

Schools in the least disadvantaged areas had higher odds of implementing *‘School provides end of trip facilities to encourage active school travel’* (85% vs. 66%; OR 2.97 (95%CI 1.03 to 8.58)) compared to schools in the most disadvantaged areas.

## Discussion

This study assessed the current implementation of various physical activity initiatives in Australian primary schools and their association with school characteristics. Implementation rates varied considerably, ranging between 4% and 98% across the four opportunities for physical activity. Of the 33 initiatives examined, 15 were implemented in over 50% of schools and only four in more than 75%. Most initiatives showed similar implementation rates across the geographic characteristics, with only nine significantly associated with school size, location, or socio-economic status. Specifically, smaller and larger schools, urban schools, and less disadvantaged schools were more likely to implement certain physical activity initiatives.

National data on the implementation of physical activity initiatives in Australian primary schools is very limited, hindering meaningful comparisons. The only national data comes from the Active Healthy Kids Australia 2022 report card which showed that only 43% to 50% of teachers nationally rated their facilities and equipment available for PE and sport as good or excellent.^4^ While our study included items that measured school facilities and equipment, a limitation of the national survey is that we measured prevalence and did not collect data on the fidelity or quality of implementation.

Our study’s results generally align with state-level data on physical activity initiatives in primary schools. A 2017 study of 2033 NSW primary schools reported that 91% encouraged physical activity during recess and lunch (e.g. playground markings),^36^ while we found that 96% had access to playground markings and play areas, and 86% had sports equipment during these times. Additionally, a 2021 trial in 115 NSW schools similarly showed that about half of primary schools implemented classroom active breaks (53% versus 48%).^22^ However, several differences emerged. Our study reported a higher proportion of Australian primary schools integrating physical activity into key learning areas beyond PDHPE (80%) compared to 59% in a 2013 NSW study.^21^ Conversely, we reported lower implementation rates for implementing fundamental movement skills in the PDHPE curriculum (75%), physical activity homework (18%) and written physical activity plans (29%), while earlier NSW studies reported 87%,^21^ 75%^22^ and 50%^21^ respectively. Additionally, 63% of our schools provided information to parents about physical activity, compared to 41%^22^ and 82%^21^ reported in previous studies. The reasons for these differences in the implementation are unclear and may be influenced by various factors, including differences in study samples (Australia versus NSW), data collection timing (2022-2023 versus 2013, 2017 and 2021), participant types (principals versus teachers), and the survey items and definitions used for implementation. Furthermore, one NSW study conducted within a research trial context included targeted physical activity initiatives and support (such as program resources and mentorship) that may have influenced adoption rates. For instance, physical activity homework was one of three curricular components schools were asked to implement during this trial.^22^ Lastly, the increased focus on being physically active in other key learning areas since the 2013 NSW study may also explain the increased implementation rates.

To our knowledge, this is the first study to explore the associations between the national implementation of physical activity initiatives and school characteristics, making direct comparisons with existing research challenging. While previous state-based studies in Australian primary school studies report no significant differences in implementation by characteristics,^21, 36^ our findings suggest some variations that align with broader research. For example, large urban schools showed higher odds of implementing *‘Incentives for being physically active (e*.*g. sports equipment)’*, which may be due to greater resources and infrastructure in urban areas.^37, 38^ Furthermore, urban and less disadvantaged schools had higher odds of implementing both active travel initiatives. This may be linked to factors such as shorter distances to schools,^39, 40^ better road safety^39–41^ and a higher likelihood of children in denser, urban environments engaging in active school travel,^42^ with socio-economic status also positively associated with active school travel to school.^41^ In contrast, smaller, rural schools often face higher costs and operational challenges, limiting their ability to offer diverse programs,^37, 38^ which may explain why larger schools had higher odds of implementing *‘Classroom-based programs or strategies on student motivation, goal setting or monitoring of physical activity’*. Interestingly, smaller schools had higher odds of implementing *‘Physical activity programs with an interactive family component’* than larger schools. These higher rates for family-focused initiatives may stem from findings from previous research suggesting greater family involvement in smaller schools^43, 44^ and a decrease in parent involvement as school size increases.^43^

This is the first nationally representative study in Australia, including participation from all states and school sectors, to report on the implementation of a comprehensive range of physical activity initiatives in primary schools. These initiatives were identified based on global evidence of recommended evidence-based school programs. The survey items used to assess implementation were either existing validated items or developed through consensus among a research team with considerable expertise in school-based physical activity initiatives.

Despite its strengths, several limitations should be considered when interpreting our study results. First, the reliance on principal self-reports to assess physical activity initiatives may be less accurate than teacher self-reports or observational audits, particularly for classroom-based initiatives. However, principal self-report has been shown to be valid and reliable for reporting on physical activity initiatives implemented in their school.^45^ Second, the low participation rate of about 20% among Australian primary schools may limit the generalisability of the findings; however, this is consistent with other recent Australian school-based cross-sectional surveys. Nevertheless, participating schools were similar to Australian primary schools, indicating a successful sampling methodology for a nationally representative sample. Third, the survey was conducted after the COVID-19 pandemic, which affected school operations and infrastructure, though we did not assess this impact. Additionally, due to research approval limitations, only one of the estimated 161 NSW government schools participated resulting in the use of post-stratification weighting based on state and sector, with most NSW data coming from Catholic and Independent schools. Fourth, while our survey assessed the implementation of physical activity initiatives, we did not assess the quality or fidelity of implementation of these initiatives for example, PE lesson quality has been assessed previously using the Supportive, Active, Autonomous, Fair and Enjoyable (SAAFE) teaching principle.^46^ Finally, while the physical activity initiatives assessed were based on global evidence of school-based studies, limited research has isolated the effectiveness of these discrete physical activity initiatives.

This study has important implications for policy and practice. The study identifies a range of physical activity initiatives that, despite being identified from high-quality systematic reviews and aligned with global policies and guidelines, are not widely implemented in Australia. For example, while systematic review evidence supports the benefits of increasing the quality of existing PE classes,^47^ increasing physical activity outside of the class,^48^ and incorporating short active class breaks,^49–51^ the current implementation of these initiatives remains suboptimal (34% to 53%). Active class breaks are particularly valuable, improving student focus and on-task behaviour,^52^ and cognitive function.^53^ These findings suggest that governments and support agencies should consider prioritising funding to enhance the implementation of these initiatives to reach all schools. If we aim to implement these programs at scale, we need to support schools in overcoming barriers to their adoption.^54^ The application of implementation science allows us to purposefully select strategies that address common barriers, such as teacher turnover, lack of confidence and competence among staff, and limited resources. For example, studies have shown that when these barriers are effectively addressed, schools’ compliance with physical activity initiatives significantly increases.^55–58^ Additionally, while our study offers valuable insights into current implementation, there is a need for ongoing surveillance systems to monitor and evaluate the effectiveness of these initiatives over time.^21^ These systems can help tailor implementation support to address barriers as they arise.^54^

## Data Availability

All data produced in the present work are contained in the manuscript.

## Acknowledgments

We would like to acknowledge the Australian Council for Education Research (ACER) for conducting the sample design, the researchers within the National Centre of Implementation Science (NCOIS), school staff from the participating primary schools and staff who conducted the computer assisted telephone interviews.

