## Appendices for "Prevalence of Physical Activity Initiatives in Australian Primary Schools: A Cross-Sectional Survey"

#### **Appendix A:** Ethics approvals

| Jurisdiction | Approval number |
|---|---|
| ACT Department of Education | RES 2314 |
| ACT/NSW Archdiocese of Canberra and Goulburn | NA |
| NSW Department of Education (SERAP) | 2021187 |
| Diocese of Bathurst | NA |
| Diocese of Maitland-Newcastle | NA |
| Diocese of Parramatta | NA |
| Diocese of Wagga Wagga | NA |
| NT Department of Education | 20817 |
| NT Catholic schools | NA |
| QLD Department of Education | 550/27/2550 |
| Archdiocese of Brisbane | 502 |
| Diocese of Cairns | NA |
| Diocese of Rockhampton | NA |
| Diocese of Townsville | 2021-13 |
| SA Department of Education | 2021-0074 |
| SA Catholic schools | 202134 |
| TAS Department of Education | 2022-03 |
| Archdiocese of Hobart | NA |
| VIC Department of Education | 2022_004563 |
| Archdiocese of Melbourne | 1173 |
| Diocese of Ballarat | NA |
| Diocese of Sale | NA |
| Diocese of Sandhurst | NA |
| WA Department of Education | D23/1411981 |
| Catholic Education Western Australia (four Dioceses (Broome, Bunbury, Geraldton, Perth) | RP2021/44 |

ACT: Australian Capital Territory; QLD: Queensland; NA: Not Applicable; NSW: New South Wales; NT: Northern Territory; SA: South Australia; SD: standard deviation; SES: Socio-Economic Status; SERAP: State Education Research Application Process; TAS: Tasmania; VIC: Victoria; WA: Western Australia

**Appendix B:** Physical activity initiative definitions

| Opportunities for physical activity | Recommended strategies | Initiatives | Definitions |
|---|---|---|---|
| Physical activity in the classroom | Provide students information they need to lead a physically active life | Physical activity units of work in PDHPE/HPE curriculum across all year groups | Units of work for teaching physical activity in the PDHPE/HPE curriculum were included across all year groups |
| | | Sedentary behaviour units of work in PDHPE/HPE curriculum across all year groups | Units of work for teaching sedentary behaviour in the PDHPE/HPE curriculum were included across all year groups |
| | | Classroom-based programs or strategies on student motivation, goal setting or monitoring of physical activity | Classroom-based programs and strategies targeting student motivation, goal setting or self-monitoring of physical activity implemented in the last 12 months |
| | | Physical activity homework provided to students at least once a week to all year groups | Physical activity homework provided at least once a week to all year groups in the last 12 months |
| | Deliver quality, active and engaging PE | Strategies to increase the frequency or duration of PE classes, physical activity or sport across all year groups | Strategies to increase the frequency or duration of PE classes, physical activity or sports were implemented across all year groups in the last 12 months |
| | | Programs to increase the quality of PE classes across all year groups | The school has implemented strategies to increase the intensity or quality of PE across all year groups in the last 12 months |
| | | Fundamental movement skills units of work in PDHPE/HPE curriculum across all year groups | Units of work for teaching fundamental movement skills in the PDHPE/HPE curriculum were included across all year groups. |
| | | Some students provided physical activity trackers | Physical activity trackers were provided to some students in the last 12 months |
| | | Teachers acted as physical activity role models for students | Teachers have been acknowledged or role modelled physical activity in the last 12 months |
| | | Active video games used in the majority of classrooms at least once a week | Active video games are used in the majority of classrooms at least once a week |
| | Provide opportunities for children to break up sitting time | Active class breaks in ≥80% of classes daily (e.g. energisers) | Physical activity breaks are implemented in greater than 80% of classes daily (e.g. energisers) |
| | | Physical activity integrated into at least some KLAs other than PDHPE | Physical activity integrated into at least some KLAs other than PDHPE |
| | | Standing desks available to at least some students in some classrooms | Standing desks are available to at least some students in some classrooms |
| Physical activity outside the classroom and during break times | Encourage physical activity and active play during recess and lunch breaks | Students are allowed to wear existing sports uniforms everyday | All students can wear existing sports uniforms, including joggers everyday |
| | | School infrastructure that supports physical activity during breaks everyday (e.g. play areas) | Students have access to play areas or fields, playground markings or outdoor play equipment 5 days per week at lunch and recess. |
| | | Sports equipment that supports physical activity during breaks everyday (e.g. basketballs) | Students have access to sports equipment 5 days per week at lunch and recess. |
| | | Teacher-led physical activity during breaks at least once a week | Teachers led organised physical activity for students at recess or lunch at least once per week |
| | | Student-led physical activity during breaks at least once a week | Student led organised physical activity at recess or lunch at least once per week |
| | Provide opportunities for children to engage in | Physical activity programs outside of class across all year groups | Programs or strategies to increase students' physical activity outside of class time across all year groups in the last 12 months |
| | | Incentives for being physically active (e.g. sports equipment) | The school has implemented a reward or incentive system to acknowledge physical activity in the last 12 months (e.g. sports equipment, water bottles, a merit system) |

| Opportunities for physical activity | Recommended strategies | Initiatives | Definitions |
|---|---|---|---|
| | physical activity outside of class time | School-wide physical activity programs (e.g. jump rope for heart) | The school has participated in school-wide programs that promoted messages about physical activity in the last 12 months (e.g. jump rope for heart) |
| | | Students supported to be physical activity role models for peers | Students have been acknowledged as, or supported to become physical activity role models in the last 12 months |
| | | Community members acted as physical activity role models for students | The school has promoted or engaged a community member as a physical activity role model in the last 12 months |
| Physical activity outside of school or involving families | Support children to be active outside of school | Information to parents to encourage active school travel | School provides information to parents to encourage safe active school travel |
| | | School provides end of trip facilities to encourage active school travel | School provides end of trip facilities to encourage active school travel |
| | | Physical activity programs delivered in partnership with community organisations or services | The school partnered with community organisations or services to deliver physical activity programs in the last 12 months |
| | | Physical activity programs with an interactive family component | Physical activity programs with an interactive family component delivered to students in the last 12 months |
| | | Information on physical activity sent home to parents (e.g. strategies to increase physical activity) | Physical activity information (e.g. strategies to increase physical activity) sent home to parents or carers in the last 12 months |
| | | Workshops with parents on physical activity | The school conducted workshops for parents/carers which included information on student physical activity in the last 12 months |
| Other | | Individual physical activity advice or counselling for all students (e.g. activity plans) | Individually tailored advice or counselling to improve physical activity was provided to all students in the last 12 months |
| | | The majority of teaching staff participated in physical activity professional development | The majority of teaching staff participated in physical activity professional development/learning in the last 12 months |
| | | School has a written physical activity plan or policy | The school has a written physical activity plan or policy |
| | | Information about school physical activity strategies/programs shared with the school community/education sector | The school has shared information on physical activity strategies or programs with the school community or sector in the last 12 months |

KLAs: Key Learning Areas; PDHPE/HPE: Personal Development, Health and Physical Education; PE: Physical Education

**Appendix C:** Principal survey

| Survey item | Responses |
|---|---|
| **School and principal characteristics** | |
| What primary school Year groups does your school cater for? | All = All of the below: Kindergarten/Prep/Reception/Pre-primary year to Year 6<br>K = Kindergarten/Prep/Reception/Pre-primary<br>1 = Year 1<br>2 = Year 2<br>3 = Year 3<br>4 = Year 4<br>5 = Year 5<br>6 = Year 6 |
| Which best describes your current role? | 1 = Executive Principal<br>2 = Principal<br>3 = Deputy Principal<br>4 = Assistant Principal<br>5 = Acting Principal<br>6 = Other role<br>7 = Head of school / Campus / Operations<br>8 = School counsellor / social worker<br>9 = Lead / classroom teacher<br>10 = Health and Wellbeing Teacher / Coordinator<br>11 = Wellbeing and Aboriginal Education Leader |
| What is your current role? | |
| How long have you been in this role at this school? | 1 = 12 months or more<br>2 = Less than 12 months |
| Please enter the number of years you have been in this role at this school | |
| **Physical activity module** | |
| For which Year groups does your school's PDHPE/HPE curriculum include programs with units of work for teaching physical activity? Note: Curriculum could include knowledge/understanding about the benefits of physical activity | 1 = Kindergarten/Prep/Reception/Pre-primary<br>2 = Year 1/Year 2<br>3 = Year 3/Year 4<br>4 = Year 5/Year 6<br>All = All Year groups<br>7 = None of the above<br>888 = Unsure |
| Which Year groups does your school have units of work regarding physical activity in Learning Areas/Key Learning Areas other than PDHPE/HPE? | 1 = Kindergarten/Prep/Reception/Pre-primary<br>2 = Year 1/Year 2<br>3 = Year 3/Year 4<br>4 = Year 5/Year 6<br>All = All Year groups<br>7 = None of the above<br>888 = Unsure |
| For which Year groups does your school's PDHPE/HPE curriculum include programs with units of work for teaching about reducing sedentary behaviours or screen time? | 1 = Kindergarten/Prep/Reception/Pre-primary<br>2 = Year 1/Year 2<br>3 = Year 3/Year 4<br>4 = Year 5/Year 6<br>All = All Year groups<br>7 = None of the above<br>888 = Unsure |
| For which Year groups does your schools PDHPE/HPE curriculum include programs with units of work for teaching and/or assessment of Fundamental Movement Skills? | 1 = Kindergarten/Prep/Reception/Pre-primary<br>2 = Year 1/Year 2<br>3 = Year 3/Year 4<br>4 = Year 5/Year 6<br>All = All Year groups<br>7 = None of the above<br>888 = Unsure |
| For which Year groups has your school implemented programs or strategies to increase the frequency or duration of PE classes, physical activity or sport in the last 12 months? | 1 = Kindergarten/Prep/Reception/Pre-primary<br>2 = Year 1/Year 2<br>3 = Year 3/Year 4<br>4 = Year 5/Year 6<br>All = All Year groups<br>7 = None of the above<br>888 = Unsure |

| Survey item | Responses |
|---|---|
| Please enter the program name(s) or a brief description of the strategies used. Note: Please separate program name(s)/strategies with a comma | |
| For which Year groups has your school implemented programs or strategies to increase the intensity or quality of PE in the last 12 months? | 1 = Kindergarten/Prep/Reception/Pre-primary<br>2 = Year 1/Year 2<br>3 = Year 3/Year 4<br>4 = Year 5/Year 6<br>All = All Year groups<br>7 = None of the above<br>888 = Unsure |
| Please enter the program name(s) or a brief description of the strategies used. Note: Please separate program name(s)/strategies with a comma | |
| For which Year groups has your school implemented programs or strategies to increase student physical activity outside of class time in the last 12 months? | 1 = Kindergarten/Prep/Reception/Pre-primary<br>2 = Year 1/Year 2<br>3 = Year 3/Year 4<br>4 = Year 5/Year 6<br>All = All Year groups<br>7 = None of the above<br>888 = Unsure |
| Please enter the program name(s) or a brief description of the strategies used. Note: Please separate program name(s)/strategies with a comma | |
| Does your school have any of the following that all students can access every day at recess and/or lunch? | 1 = Large asphalt areas suitable for games e.g., basketball/netball - recess<br>2 = Large asphalt areas suitable for games e.g., basketball/netball - lunch<br>3 = Large playing fields suitable for games e.g., soccer, football - recess<br>4 = Large playing fields suitable for games e.g., soccer, football - lunch<br>5 = Indoor activity spaces e.g., multi-purpose rooms and gymnasiums - recess<br>6 = Indoor activity spaces e.g., multi-purpose rooms and gymnasiums - lunch<br>7 = Playground markings e.g., hopscotch, wall targets - recess<br>8 = Playground markings e.g., hopscotch, wall targets - lunch<br>9 = Fixed playground equipment e.g., climbing structures, swings - recess<br>10 = Fixed playground equipment e.g. climbing structures, swings - lunch<br>11 = Sports equipment provided by the school e.g., basketballs, netballs, skipping ropes - recess<br>12 = Sports equipment provided by the school e.g., basketballs, netballs, skipping ropes - lunch<br>13 = No, none of the above<br>888 = Unsure |
| In a typical week, how many days per week does your school run teacher-led organised physical activity at recess and/or lunch for students (not including sports team training)? | 1 = 1<br>2 = 2<br>3 = 3<br>4 = 4<br>5 = 5<br>6 = Less than 1 day/week<br>7 = None<br>888 = Unsure |
| In a typical week, how many days per week does your school run student-led organised physical activity at recess and/or lunch? | 1 = 1<br>2 = 2<br>3 = 3<br>4 = 4<br>5 = 5<br>6 = Less than 1 day/week<br>7 = None<br>888 = Unsure |

| Survey item | Responses |
|---|---|
| Does your school provide bike or scooter racks for students use? | 1 = Yes<br>2 = No<br>888 = Unsure |
| Do classes at your school implement active class breaks during class time? Note: These are short, structured class-based physical activity breaks e.g., energisers | 1 = Yes<br>2 = No<br>888 = Unsure |
| What percentage of all classes at your school would you estimate implement active class breaks during class time? | |
| On how many days per week do active class breaks usually occur in participating classes? | 1 = 1<br>2 = 2<br>3 = 3<br>4 = 4<br>5 = 5<br>6 = Less than 1 day/week<br>7 = None<br>888 = Unsure |
| Do classes at your school integrate physical activity into Key Learning Areas other than PDHPE/HPE? | 1 = Yes, all classes<br>2 = Yes, some classes<br>3 = No<br>888 = Unsure |
| In what proportion of classrooms are standing desks available for students to use at your school? | 1 = All classrooms<br>2 = Most classrooms (more than half but not all)<br>3 = Some classrooms (but less than half)<br>4 = No classrooms - we do not have standing desks available<br>888 = Unsure |
| For how many students are standing desks available for in these classrooms? | 1 = All students<br>2 = Most students (more than half but not all)<br>3 = Some students (but less than half)<br>888 = Unsure |
| Has your school provided students with physical activity trackers to wear in PE or any other time in the last 12 months? Note: Physical activity trackers could include pedometers, activity watches | 1 = Yes, to all students<br>2 = Yes, to selected students (e.g., some classes)<br>3 = Yes, as part of a research trial<br>4 = No<br>888 = Unsure |
| In what proportion of classes are active video games used with students during class time? Note: This could include Just Dance | 1 = All classrooms<br>2 = Most classrooms (more than half but not all)<br>3 = Some classrooms (but less than half)<br>4 = No classrooms - we do not have active video games available<br>888 = Unsure |
| In classes where active video games are used, how frequently are they used? | 1 = Daily<br>2 = At least weekly<br>3 = At least monthly<br>4 = Once a term or less frequently<br>888 = Unsure |
| Has your school implemented any physical activity programs or strategies for students which include an interactive parent/family/carer in the last 12 months? Note: This could include parent/child workshops that involve active participation or physical activity classes | 1 = Yes<br>2 = No<br>888 = Unsure |
| Please enter the program name(s) or a brief description of the strategies with interactive. Note: Please separate program name(s)/strategies with a comma | |
| Does your school allow all students to wear your schools existing sports uniform, including joggers, every day of the week? | 1 = Yes, all students can wear the sports uniform every day<br>2 = No, sports uniform is only allowed on designated days/time with a separate traditional uniform at all other times<br>888 = Unsure |
| Has your school implemented any reward or incentive systems to acknowledge student physical activity, in the last 12 months? Note: Small rewards or incentives could include sports equipment, water bottles, a merit system | 1 = Yes<br>2 = No<br>888 = Unsure |
| Please describe any reward or incentive systems that acknowledged student physical activity, in the last 12 months | |

| Survey item | Responses |
|---|---|
| Are there any other programs or strategies that your school implements targeting physical activity that we haven't asked you specifically about? | 1 = Yes<br>2 = No<br>888 = Unsure |
| Please describe the other programs or strategies that your school implements which targets physical activity | |
| **Physical activity and healthy eating module** | |
| For which Year groups has your school provided students with weekly healthy eating and/or physical activity homework in the last 12 months? | 1 = Kindergarten/Prep/Reception/Pre-primary – healthy eating<br>2 = Kindergarten/Prep/Reception/Pre-primary – physical activity<br>3 = Year 1/Year 2 - healthy eating<br>4 = Year 1/Year 2 - physical activity<br>5 = Year 3/Year 4 - healthy eating<br>6 = Year 3/Year 4 - physical activity<br>7 = Year 5/Year 6 - healthy eating<br>8 = Year 5/Year 6 - physical activity<br>9 = None of the above<br>888 = Unsure |
| Has your school sent information home to parents/carers about any of the following healthy eating and/or physical activity topics in the last 12 months? Note: This could include via letter, newsletter, pamphlet, email, website or parent meetings | 1 = Healthy lunchboxes<br>2 = Healthy canteen<br>3 = Vegetable, fruit and water breaks<br>4 = Benefits of, or strategies to increase physical activity<br>5 = School policy or guidelines regarding healthy eating<br>6 = Limiting small screen recreation<br>7 = Benefits of safe active travel<br>8 = Encouragement to 'park and walk' with their children to school<br>9 = School policy or guideline regarding physical activity<br>10 = Other<br>11 = None of the above<br>12 = Benefits of, or strategies to increase healthy eating or water consumption<br>888 = Unsure |
| What other healthy eating and/or physical activity topics has your school sent information home to parents/carers about? | |
| Has your school conducted any workshops for parents/carers (either face-to-face or online) which included information on student healthy eating and/or physical activity in the last 12 months? | 1 = Yes, safe walking, cycling, scootering or other active travel activities<br>2 = Yes, strategies to improve student healthy eating<br>3 = Yes, strategies to improve student physical activity<br>4 = Yes, other<br>5 = No, did not conduct any workshops<br>6 = yes, strategies to improve general wellbeing including healthy eating and physical activity<br>888 = Unsure |
| Please describe the other workshops for parents/carers | |
| What proportion of your teaching staff have participated in physical activity professional development/learning in the last 12 months? | 1 = All staff<br>2 = Most staff (more than half but not all)<br>3 = Some staff (but less than half)<br>4 = No staff<br>888 = Unsure |
| Does your school have a written physical activity plan or policy? Note: This can include a general policy that explicitly references physical activity | 1 = Yes<br>2 = No<br>888 = Unsure |
| Has your school shared information on your healthy eating and/or physical activity strategies or programs with your school community or education sector in the last 12 months? Note: School community or education sector could include a School Board, P&C, or school education director/principal/region director/executive group | 1 = Yes, healthy eating<br>2 = Yes, physical activity<br>3 = No, neither<br>888 = Unsure |
| Has your school provided students with individually tailored counselling or advice to increase their healthy eating and/or physical activity in the last 12 months? Note: This could include online programs, activity plans developed with teachers | 1 = Yes, heathy eating to all students<br>2 = Yes, healthy eating to selected students<br>3 = Yes, physical activity to all students<br>4 = Yes, physical activity to selected students<br>5 = None of the above<br>888 = Unsure |

| Survey item | Responses |
|---|---|
| Has your school participated in any school-wide initiatives that promoted messages about healthy eating and/or physical activity in the last 12 months? (e.g., Fruit and Vege month, Jump Rope for Heart). Note: Ways of promoting messages school-wide could include putting up posters around school, hosting school events - this does not include strategies that only included providing information to parents | 1 = Yes, healthy eating<br>2 = Yes, physical activity<br>3 = No, neither<br>888 = Unsure |
| Have any students at your school been acknowledged as, or supported to become healthy eating and/or physical activity role models in the last 12 months? | 1 = Yes, healthy eating<br>2 = Yes, physical activity<br>3 = No, neither<br>888 = Unsure |
| Have any staff members at your school been acknowledged as, or acted as healthy eating and/or physical activity role models in the last 12 months? | 1 = Yes, healthy eating<br>2 = Yes, physical activity<br>3 = No, neither<br>888 = Unsure |
| Has your school promoted or engaged any people outside of your school community as healthy eating and/or physical activity role models in the last 12 months? (e.g., local or well-known athlete) | 1 = Yes, healthy eating<br>2 = Yes, physical activity<br>3 = No, neither<br>888 = Unsure |
| Has your school partnered with any community organisations or services to deliver programs targeting student healthy eating and/or physical activity in the last 12 months? Note: Community organisations or services could include working with local sports clubs or local community consultation to develop programs | 1 = Yes, healthy eating<br>2 = Yes, physical activity<br>3 = No, neither<br>888 = Unsure |
| Has your school implemented any classroom-based programs or strategies targeting student's motivation, personal goals or self-monitoring of healthy eating and/or physical activity in the last 12 months? Note: Classroom-based programs or strategies could include activity plans, journals either in class or at home | 1 = Yes, healthy eating<br>2 = Yes, physical activity<br>3 = No, neither<br>888 = Unsure |
| Are there any other programs or strategies that your school implements targeting healthy eating or physical activity that we haven't asked you specifically about? | 1 = Yes<br>2 = No<br>888 = Unsure |
| Please describe the other programs or strategies that your school implements which targets healthy eating or physical activity | |

PDHPE/HPE: Personal Development, Health and Physical Education; PE: Physical Education

### **Appendix D:** Summary of out-of-scope schools

| Reason for being out of scope | n |
|---|---|
| Taking part in other trial | 210 |
| Not invited due to research approval not being extended (NSW only) | 846 |
| School closed / no current staff or students | 5 |
| Distance schools | 2 |
| Does not accept phone calls | 1 |

NSW: New South Wales

**Appendix E.** Associations between implementation of physical activity initiatives and school characteristics

| Initiatives | School characteristic | Category | Prevalence of implementation n (%) | Simple regression | | |
|---|---|---|---|---|---|---|
| | | | | OR (95% CI) | P value | N |
| Physical activity units of work in PDHPE/HPE curriculum across all year groups | School size | Small | 180 (96.65) | 0.40 (0.07;2.22) | 0.30 | 289 |
| | | Large | 98 (98.62) | | | |
| | Remoteness | Rural areas | 159 (95.93) | 0.20 (0.03 to 1.46) | 0.11 | |
| | | Urban areas | 119 (99.18) | | | |
| | SES | Least Disadvantaged | 108 (97.81) | 1.20 (0.27 to 5.24) | 0.81 | |
| | | Most Disadvantaged | 170 (97.39) | | | |
| Sedentary behaviour units of work in PDHPE/HPE curriculum across all year groups | School size | Small | 84 (44.72) | 0.48 (0.23 to 0.98) | **0.04**[a] | 288 |
| | | Large | 48 62.99) | | | |
| | Remoteness | Rural areas | 73 (45.24) | 0.52 (0.26 to 1.06) | 0.07 | |
| | | Urban areas | 59 (61.19) | | | |
| | SES | Least Disadvantaged | 55 (61.70) | 1.88 (0.88 to 3.99) | 0.10 | |
| | | Most Disadvantaged | 77 (46.16) | | | |
| Classroom-based programs strategies on student motivation, goal setting monitoring of physical activity | School size | Small | 40 (22.90) | 0.44 (0.20 to 0.93) | **0.03**[a] | 267 |
| | | Large | 31 (40.45) | | | |
| | Remoteness | Rural areas | 37 (23.70) | 0.50 (0.23 to 1.07) | 0.07 | |
| | | Urban areas | 34 (38.53) | | | |
| | SES | Least Disadvantaged | 27 30.51) | 0.94 (0.43 to 2.04) | 0.87 | |
| | | Most Disadvantaged | 44 (31.85) | | | |
| Physical activity homework provided to students at least once a week to all year groups | School size | Small | 30 (15.66) | 0.68 (0.30 to 1.54) | 0.36 | 268 |
| | | Large | 17 (21.34) | | | |
| | Remoteness | Rural areas | 28 (17.38) | 0.88 (0.39 to 1.98) | 0.75 | |
| | | Urban areas | 19 (19.32) | | | |
| | SES | Least Disadvantaged | 14 (15.76) | 0.73 (0.30 to 1.78) | 0.49 | |
| | | Most Disadvantaged | 33 (20.35) | | | |
| Strategies to increase the frequency or duration of PE classes, physical activity or sport across all year groups | School size | Small | 114 (57.41) | 1.61 (0.72 to 3.60) | 0.25 | 287 |
| | | Large | 38 (45.58) | | | |
| | Remoteness | Rural areas | 98 (56.99) | 1.50 (0.70 to 3.23) | 0.29 | |
| | | Urban areas | 54 (46.83) | | | |
| | SES | Least Disadvantaged | 51 (44.07) | 0.56 (0.27 to 1.15) | 0.12 | |
| | | Most Disadvantaged | 101 (58.51) | | | |
| Programs to increase the quality of PE classes across all year groups | School size | Small | 80 (41.86) | 2.23 (1.05 to 4.73) | **0.04**[a] | 285 |
| | | Large | 31 (24.44) | | | |
| | Remoteness | Rural areas | 69 (37.92) | 1.46 (0.71 to 3.00) | 0.30 | |
| | | Urban areas | 42 (29.46) | | | |
| | SES | Least Disadvantaged | 39 (33.26) | 0.97 (0.47 to 2.01) | 0.94 | |
| | | Most Disadvantaged | 72 (33.90) | | | |
| Fundamental movement skills units of work in PDHPE/HPE curriculum across all year groups | School size | Small | 142 (77.84) | 1.44 (0.69 to 3.02) | 0.33 | 287 |
| | | Large | 63 (70.89) | | | |
| | Remoteness | Rural areas | 122 (79.29) | 1.64 (0.80 to 3.37) | 0.17 | |

| Initiatives | School characteristic | Category | Prevalence of implementation n (%) | Simple regression | | |
|---|---|---|---|---|---|---|
| | | | | OR (95% CI) | P value | N |
| | | Urban areas | 83 (69.98) | | | |
| | SES | Least Disadvantaged | 75 (73.56) | 0.91 (0.44 to 1.86) | 0.79 | |
| | | Most Disadvantaged | 130 (75.43) | | | |
| Some students provided physical activity trackers | School size | Small | 9 (4.19) | 0.91 (0.21 to 4.07) | 0.91 | 282 |
| | | Large | 3 (4.56) | | | |
| | Remoteness | Rural areas | 7 (3.61) | 0.70 (0.17 to 2.79) | 0.61 | |
| | | Urban areas | 5 (5.10) | | | |
| | SES | Least Disadvantaged | 2 (3.25) | 0.60 (0.10 to 3.48) | 0.57 | |
| | | Most Disadvantaged | 10 (5.31) | | | |
| Teachers acted as physical activity role models for students | School size | Small | 32 (15.56) | 0.86 (0.34 to 2.16) | 0.74 | 268 |
| | | Large | 17 (17.71) | | | |
| | Remoteness | Rural areas | 25 (11.21) | 0.45 (0.18 to 1.14) | 0.09 | |
| | | Urban areas | 24 (21.74) | | | |
| | SES | Least Disadvantaged | 20 (19.05) | 1.36 (0.54 to 3.46) | 0.52 | |
| | | Most Disadvantaged | 29 (14.73) | | | |
| Active video games used in the majority of classrooms at least once a week | School size | Small | 18 (7.01) | 1.00 (0.30 to 3.28) | 1.00 | 282 |
| | | Large | 9 (7.03) | | | |
| | Remoteness | Rural areas | 13 (5.43) | 0.61 (0.20 to 1.84) | 0.38 | |
| | | Urban areas | 14 (8.55) | | | |
| | SES | Least Disadvantaged | 13 (10.47) | 2.74 (0.97 to 7.78) | 0.058 | |
| | | Most Disadvantaged | 14 (4.09) | | | |
| Active class breaks in ≥80% of classes daily (e.g. energisers) | School size | Small | 90 (44.81) | 0.51 (0.24 to 1.05) | 0.07 | 282 |
| | | Large | 45 (61.61) | | | |
| | Remoteness | Rural areas | 77 (47.32) | 0.65 (0.31 to 1.35) | 0.25 | |
| | | Urban areas | 58 (57.98) | | | |
| | SES | Least Disadvantaged | 48 (54.54) | 1.14 (0.54 to 2.42) | 0.73 | |
| | | Most Disadvantaged | 87 (51.22) | | | |
| Physical activity integrated into at least some KLAs other than PDHPE | School size | Small | 144 (79.71) | 0.94 (0.43 to 2.06) | 0.88 | 282 |
| | | Large | 70 (80.63) | | | |
| | Remoteness | Rural areas | 126 (77.60) | 0.73 (0.34 to 1.57) | 0.42 | |
| | | Urban areas | 88 (82.59) | | | |
| | SES | Least Disadvantaged | 83 (85.57) | 1.92 (0.88 to 4.17) | 0.10 | |
| | | Most Disadvantaged | 131 (75.53) | | | |
| Standing desks available to at least some students in some classrooms | School size | Small | 67 (31.70) | 0.50 (0.23 to 1.11) | 0.09 | 282 |
| | | Large | 52 (47.92) | | | |
| | Remoteness | Rural areas | 59 (33.96) | 0.64 (0.30 to 1.38) | 0.25 | |
| | | Urban areas | 60 (44.59) | | | |
| | SES | Least Disadvantaged | 47 (47.30) | 1.85 (0.89 to 3.86) | 0.10 | |
| | | Most Disadvantaged | 72 (32.62) | | | |
| Students are allowed to wear existing sports uniforms everyday | School size | Small | 120 (62.25) | 0.75 (0.33 to 1.70) | 0.49 | 282 |
| | | Large | 54 (68.81) | | | |

| Initiatives | School characteristic | Category | Prevalence of implementation n (%) | Simple regression | | |
|---|---|---|---|---|---|---|
| | | | | OR (95% CI) | P value | N |
| | Remoteness | Rural areas | 115 (67.90) | 1.25 (0.57 to 2.74) | 0.58 | |
| | | Urban areas | 59 (62.88) | | | |
| | SES | Least Disadvantaged | 63 (59.64) | 0.63 (0.29 to 1.35) | 0.24 | |
| | | Most Disadvantaged | 111 (70.19) | | | |
| School infrastructure that supports physical activity during breaks everyday (e.g. play areas) | School size | Small | 176 (94.79) | 0.32 (0.04 to 2.76) | 0.30 | 283 |
| | | Large | 96 (98.27) | | | |
| | Remoteness | Rural areas | 157 (95.55) | 0.60 (0.11 to 3.25) | 0.55 | |
| | | Urban areas | 115 (97.29) | | | |
| | SES | Least Disadvantaged | 103 (95.90) | 0.75 (0.16 to 3.50) | 0.72 | |
| | | Most Disadvantaged | 169 (96.88) | | | |
| Sports equipment that supports physical activity during breaks everyday (e.g. basketballs) | School size | Small | 168 (88.46) | 1.50 (0.56 to 4.02) | 0.42 | 283 |
| | | Large | 86 (83.66) | | | |
| | Remoteness | Rural areas | 150 (87.66) | 1.28 (0.48 to 3.42) | 0.63 | |
| | | Urban areas | 104 (84.77) | | | |
| | SES | Least Disadvantaged | 94 (81.92) | 0.51 (0.20 to 1.34) | 0.17 | |
| | | Most Disadvantaged | 160 (89.81) | | | |
| Teacher-led physical activity during breaks at least once a week | School size | Small | 95 (50.83) | 0.90 (0.40 to 2.01) | 0.79 | 282 |
| | | Large | 58 (53.52) | | | |
| | Remoteness | Rural areas | 79 (48.50) | 0.75 (0.34 to 1.65) | 0.48 | |
| | | Urban areas | 69 (55.57) | | | |
| | SES | Least Disadvantaged | 56 (57.79) | 1.53 (0.73 to 3.22) | 0.26 | |
| | | Most Disadvantaged | 92 (47.26) | | | |
| Student-led physical activity during breaks at least once a week | School size | Small | 90 (45.05) | 0.86 (0.39 to 1.91) | 0.72 | 282 |
| | | Large | 47 (48.68) | | | |
| | Remoteness | Rural areas | 83 (48.59) | 1.15 (0.53 to 2.53) | 0.72 | |
| | | Urban areas | 54 (45.00) | | | |
| | SES | Least Disadvantaged | 47 (38.34) | 0.53 (0.25 to 1.12) | 0.09 | |
| | | Most Disadvantaged | 90 (53.92) | | | |
| Physical activity programs outside of class across all year groups | School size | Small | 86 (46.05) | 0.71 (0.33 to 1.50) | 0.37 | 284 |
| | | Large | 42 (54.69) | | | |
| | Remoteness | Rural areas | 73 (40.57) | 0.47 (0.23 to 0.96) | **0.04**[a] | |
| | | Urban areas | 55 (59.33) | | | |
| | SES | Least Disadvantaged | 45 (48.61) | 0.89 (0.42 to 1.89) | 0.76 | |
| | | Most Disadvantaged | 83 (51.48) | | | |
| Incentives for being physically active (e.g. sports equipment) | School size | Small | 42 (22.70) | 0.39 (0.16 to 0.93) | **0.03**[a] | 282 |
| | | Large | 31 (42.90) | | | |
| | Remoteness | Rural areas | 36 (21.66) | 0.37 (0.16 to 0.87) | **0.02**[a] | |
| | | Urban areas | 37 (42.46) | | | |
| | SES | Least Disadvantaged | 30 (33.11) | 1.08 (0.42 to 2.73) | 0.88 | |
| | | Most Disadvantaged | 43 (31.50) | | | |
| | School size | Small | 96 (61.02) | 1.10 (0.53 to 2.26) | 0.80 | 268 |

| Initiatives | School characteristic | Category | Prevalence of implementation n (%) | Simple regression | | |
|---|---|---|---|---|---|---|
| | | | | OR (95% CI) | P value | N |
| School-wide physical activity programs (e.g. jump rope for heart) | | Large | 50 (58.79) | | | |
| | Remoteness | Rural areas | 87 (61.25) | 1.11 (0.55 to 2.26) | 0.77 | |
| | | Urban areas | 59 (58.72) | | | |
| | SES | Least Disadvantaged | 59 (58.16) | 0.88 (0.42 to 1.82) | 0.72 | |
| | | Most Disadvantaged | 87 (61.31) | | | |
| Students supported to be physical activity role models for peers | School size | Small | 29 (12.27) | 0.61 (0.25 to 1.49) | 0.28 | 268 |
| | | Large | 19 (18.60) | | | |
| | Remoteness | Rural areas | 24 (13.97) | 0.82 (0.33 to 2.03) | 0.66 | |
| | | Urban areas | 24 (16.56) | | | |
| | SES | Least Disadvantaged | 24 (17.94) | 1.43 (0.58 to 3.49) | 0.44 | |
| | | Most Disadvantaged | 24 (13.29) | | | |
| Community members acted as physical activity role models for students | School size | Small | 58 (33.81) | 0.71 (0.34 to 1.48) | 0.36 | 268 |
| | | Large | 36 (41.91) | | | |
| | Remoteness | Rural areas | 51 (30.25) | 0.53 (0.26 to 1.11) | 0.09 | |
| | | Urban areas | 43 (44.78) | | | |
| | SES | Least Disadvantaged | 34 (34.13) | 0.77 (0.36 to 1.62) | 0.48 | |
| | | Most Disadvantaged | 60 (40.35) | | | |
| Information to parents to encourage active school travel | School size | Small | 62 (38.78) | 0.52 (0.25 to 1.09) | 0.08 | 268 |
| | | Large | 44 (54.82) | | | |
| | Remoteness | Rural areas | 47 (32.16) | 0.31 (0.15 to 0.67) | **0.002**[a] | |
| | | Urban areas | 59 (60.10) | | | |
| | SES | Least Disadvantaged | 55 (56.82) | 2.09 (1.00 to 4.40) | 0.05 | |
| | | Most Disadvantaged | 51 (38.59) | | | |
| School provides end of trip facilities to encourage active school travel | School size | Small | 134 (70.61) | 0.63 (0.17 to 2.36) | 0.49 | 282 |
| | | Large | 88 (79.24) | | | |
| | Remoteness | Rural areas | 123 (76.48) | 1.21 (0.42 to 3.44) | 0.73 | |
| | | Urban areas | 99 (72.95) | | | |
| | SES | Least Disadvantaged | 94 (85.13) | 2.97 (1.03 to 8.58) | **0.04**[a] | |
| | | Most Disadvantaged | 128 (65.81) | | | |
| Physical activity programs delivered in partnership with community organisations or services | School size | Small | 82 (48.11) | 0.78 (0.38 to 1.59) | 0.50 | 267 |
| | | Large | 45 (54.24) | | | |
| | Remoteness | Rural areas | 73 (44.77) | 0.61 (0.30 to 1.24) | 0.17 | |
| | | Urban areas | 54 (57.03) | | | |
| | SES | Least Disadvantaged | 47 (47.78) | 0.80 (0.39 to 1.64) | 0.53 | |
| | | Most Disadvantaged | 80 (53.49) | | | |
| Physical activity programs with an interactive family component | School size | Small | 35 (19.02) | 3.30 (1.23 to 8.86) | **0.02**[a] | 282 |
| | | Large | 10 (6.64) | | | |
| | Remoteness | Rural areas | 28 (16.25) | 1.71 (0.70 to 4.21) | 0.24 | |
| | | Urban areas | 17 (10.19) | | | |
| | SES | Least Disadvantaged | 20 (13.58) | 1.07 (0.44 to 2.56) | 0.88 | |
| | | Most Disadvantaged | 25 (12.82) | | | |

| Initiatives | School characteristic | Category | Prevalence of implementation n (%) | Simple regression | | |
|---|---|---|---|---|---|---|
| | | | | OR (95% CI) | P value | N |
| Information on physical activity sent home to parents (e.g. strategies to increase physical activity) | School size | Small | 87 (52.26) | 0.37 (0.16 to 0.84) | **0.02**[a] | 268 |
| | | Large | 62 (74.89) | | | |
| | Remoteness | Rural areas | 71 (52.40) | 0.40 (0.18 to 0.89) | **0.02**[a] | |
| | | Urban areas | 78 (73.26) | | | |
| | SES | Least Disadvantaged | 65 (66.34) | 1.28 (0.61 to 2.71) | 0.52 | |
| | | Most Disadvantaged | 84 (60.58) | | | |
| Workshops with parents on physical activity | School size | Small | 11 (8.99) | 0.59 (0.18 to 1.86) | 0.37 | 268 |
| | | Large | 10 (14.43) | | | |
| | Remoteness | Rural areas | 11 (10.04) | 0.74 (0.23 to 2.37) | 0.62 | |
| | | Urban areas | 10 (13.06) | | | |
| | SES | Least Disadvantaged | 8 (7.74) | 0.50 (0.16 to 1.51) | 0.22 | |
| | | Most Disadvantaged | 13 (14.48) | | | |
| Individual physical activity advice or counselling for all students (e.g. activity plans) | School size | Small | 11 (6.72) | 0.36 (0.12 to 1.12) | 0.08 | 268 |
| | | Large | 9 (16.71) | | | |
| | Remoteness | Rural areas | 8 (6.86) | 0.39 (0.10 to 1.55) | 0.18 | |
| | | Urban areas | 12 (15.92) | | | |
| | SES | Least Disadvantaged | 8 (14.17) | 1.58 (0.46 to 5.38) | 0.46 | |
| | | Most Disadvantaged | 12 (9.47) | | | |
| The majority of teaching staff participated in physical activity professional development | School size | Small | 11 (8.30) | 0.47 (0.14 to 1.54) | 0.21 | 268 |
| | | Large | 11 (16.20) | | | |
| | Remoteness | Rural areas | 9 (8.42) | 0.50 (0.14 to 1.73) | 0.27 | |
| | | Urban areas | 13 (15.56) | | | |
| | SES | Least Disadvantaged | 8 (9.04) | 0.59 (0.19 to 1.80) | 0.36 | |
| | | Most Disadvantaged | 14 (14.36) | | | |
| School has a written physical activity plan or policy | School size | Small | 51 (31.68) | 1.38 (0.62 to 3.06) | 0.44 | 268 |
| | | Large | 22 (25.21) | | | |
| | Remoteness | Rural areas | 44 (29.95) | 1.14 (0.51 to 2.52) | 0.75 | |
| | | Urban areas | 29 (27.29) | | | |
| | SES | Least Disadvantaged | 24 (22.70) | 0.60 (0.27 to 1.32) | 0.20 | |
| | | Most Disadvantaged | 49 (33.03) | | | |
| Information about school physical activity strategies/programs shared with the school community/education sector | School size | Small | 51 (27.03) | 0.54 (0.26 to 1.16) | 0.12 | 268 |
| | | Large | 30 (40.48) | | | |
| | Remoteness | Rural areas | 41 (25.08) | 0.47 (0.22 to 1.02) | 0.06 | |
| | | Urban areas | 40 (41.45) | | | |
| | SES | Least Disadvantaged | 32 (34.39) | 1.08 (0.51 to 2.30) | 0.85 | |
| | | Most Disadvantaged | 49 (32.73) | | | |

[a]Statistically significant alpha level of 0.05 (bolded); CI: Confidence Interval; KLAs: Key Learning Areas; PDHPE/HPE: Personal Development, Health and Physical Education; PE: Physical Education; OR: Odds Ratio; SES: Socio-Economic Status; School size: Small <300, Large >=300; urban: major city, rural: inner regional, outer regional, remote
